# The Incubation Period of Severe Acute Respiratory Syndrome Coronavirus 2:A Systematic Review

**DOI:** 10.1101/2020.08.01.20164335

**Authors:** Zhiyao Li, Yu Zhang, Chenchen Wang, Liuqing Peng, Rongrong Gao, Jiarui Jing, Binzhi Ren, Jianguo Xu, Tong Wang

**Affiliations:** Department of Health Statistics and Epidemiology, School of Public Health, Shanxi Medical University, Taiyuan 030001, PR China; Shanxi Provincial Center for Disease Control and Prevention, Shanxi Provincial Key Laboratory of Major Infectious Disease Prevention, Control, Diagnosis and Treatment; State Key Laboratory of Infectious Disease Prevention and Control, National Institute for Communicable Disease Control and Prevention, Chinese Center for Disease Control and Prevention, Changping, Beijing 102206, PR China; Institute of Public Health, Nankai University, Tianjing 300350, China

**Keywords:** COVID-19, SARS-CoV-2 infection, incubation period, asymptomatic infections

## Abstract

While the novel coronavirus continues to spread worldwide, the reported incubation period has varied between studies and is imprecise due to limited data. A literature search with certain selection criteria was conducted on May 30, 2020. In total, sixty-four articles were included, and 854 individual-level data were extracted from 30 studies for pooled analysis. Of these studies, 72% of them reported a median or mean incubation period of 4-7 days, while our estimated median was 4.9 days (95% confidence interval [CI]: 4.6-5.2). However, the inclusion of 81 asymptomatic and presymptomatic patients, as well as 31 cases with incubation periods exceeding 14 days, led to our estimation of 97.5 th percentile with 19.3 days (95% CI: 17.4-21.4), beyond the currently suggested 14-day quarantine period. Therefore, we appeal to prolong the quarantine duration, especially for areas that have insufficient testing resources, to protect susceptible populations from being infected.

**Article Summary Line:** This article reviewed the COVID-19 studies involving incubation period and provided pooled estimation based on available data from these studies.The result showed that our estimated median incubation period is consistent with the estimates of formal studies but the 97.5 percentile is larger than ever on account of including a number of asymptomatic and presymptomatic patients.These finds suggested that we should properly prolong the isolation or quarantine period in order to identify more patients with longer incubation period and those without any symptoms.

## Text

Since 4 initial cases were reported on December 29, 2019 in Wuhan, Hubei Province, China (1), the world still suffers from COVID-19. It has caused 10 million infections and more than 500,000 fatalities, and continues to spread globally with 90,000 new cases each day (2). As one of the non-pharmacological interventions (NPI) to control transmission, quarantine policies are mainly based on estimates of the incubation period, namely the time between infection and symptom onset (3). These have already proved effective (4) and are conducted by most countries, including China and the United States (5-6).

However, sporadic cases seem to be inevitable, even though outbreaks can generally be controlled. Asymptomatic patients who do not have any clinical symptoms but tested positive for COVID-19 by reverse transcription polymerase chain reaction (RT-PCR) or for antibodies (5,7) can be divided into two types based on current evidence: truly asymptomatic individuals without symptoms throughout the course of the disease and presymptomatic individuals who eventually develop symptoms (8). The incubation period for the latter type usually exceeds 14 days. However, both are still infectious (9) after the currently recommended 14-day isolation period suggested by the 95th or 97.5th percentiles of previous studies (10), meaning that this strategy fails to completely separate susceptible populations from infected people.

The incubation period not only contributes to infectious disease control policymaking, but also plays a fundamental role in estimating other epidemiological parameters and statistical prediction modeling, and even helps trace potential index cases (11). Unfortunately, formal research estimating the COVID-19 incubation period has limitations due to insufficient and lowly representative samples and the exclusion of cases with missing data. In addition, patients with a short incubation period and severe symptoms tended to be included more frequently than others in the early stage of the outbreak (12,13).

Policies and other parameters based on an imprecise incubation period will bring less reliability. Thus, we performed a systematic review of studies describing the COVID-19 incubation period and performed a pooled analysis of available individual-level data extracted from studies to intuitively show the consistency and differences between studies and deliver solid estimations of the incubation period distribution based on more comprehensive data.

## Methods

### Search strategy and selection criteria

We searched “incubation period”, “COVID-19”, and their common variations (Appendix) in PubMed and Google Scholar. We also searched these terms in Chinese databases, including CNKI and Wanfang, since there were a number of studies published exclusively in Chinese. We did not include preprint platforms such as bioRxiv and medRxiv in our search plan due to their uncertainty of reliability (14), while instructive preprints were usually also published in regular journals later. Searches were completed on May 30, 2020 and citations inside each article were also identified as supplements.

First, we removed duplicates, and then excluded irrelevant, non-English, and non-Chinese research after checking titles and abstracts. Second, we reviewed the full text of each study and included studies according to whether they contained information on incubation period estimates and if not all incubation period estimates described were from citations.

### Literature and data classification

All included studies were generally divided into four categories: 1) providing only incubation period estimates without sources or citations, 2) providing only incubation period estimates from original data, 3) incubation period estimates based on original data and individual-level data, or 4) providing only individual-level data. For studies in the last two categories, the time of exposure and symptom onset for each patient as well as sex, age, and location were extracted from articles, figures, and tables by two reviewers independently. Discussions and cross-checking were performed if there were disagreements and consensus results were used to ensure consistency and precision.

Due to different measurement methods and the specific situations of each study, the collected individual-level data appeared in different forms, including single- and double-interval censored data as well as exactly observed data. Moreover, there were cases without times of exposure or symptom onset due to missing relevant details and cases that remained asymptomatic throughout the survey. Such missing data also had too little information on covariates to impute with the usual methods, including multiple imputations. Therefore, we conducted data classification and used a corresponding imputation strategy (Figure 1) and transformed all data into double-interval censored data as the optimal form to obtain the most precise estimates (15). Each patient eventually had lower and upper limits of possible exposure times and symptom onset (i.e., EL, EU, SL, and SR). Although there were patients who developed symptoms within the possible time interval of exposure, we assumed that the exposure must precede the symptoms. Thus, cases whose SU≤EU or SL≤EL were excluded and the rest of the data proceeded to further analysis.

**Figure 1:**
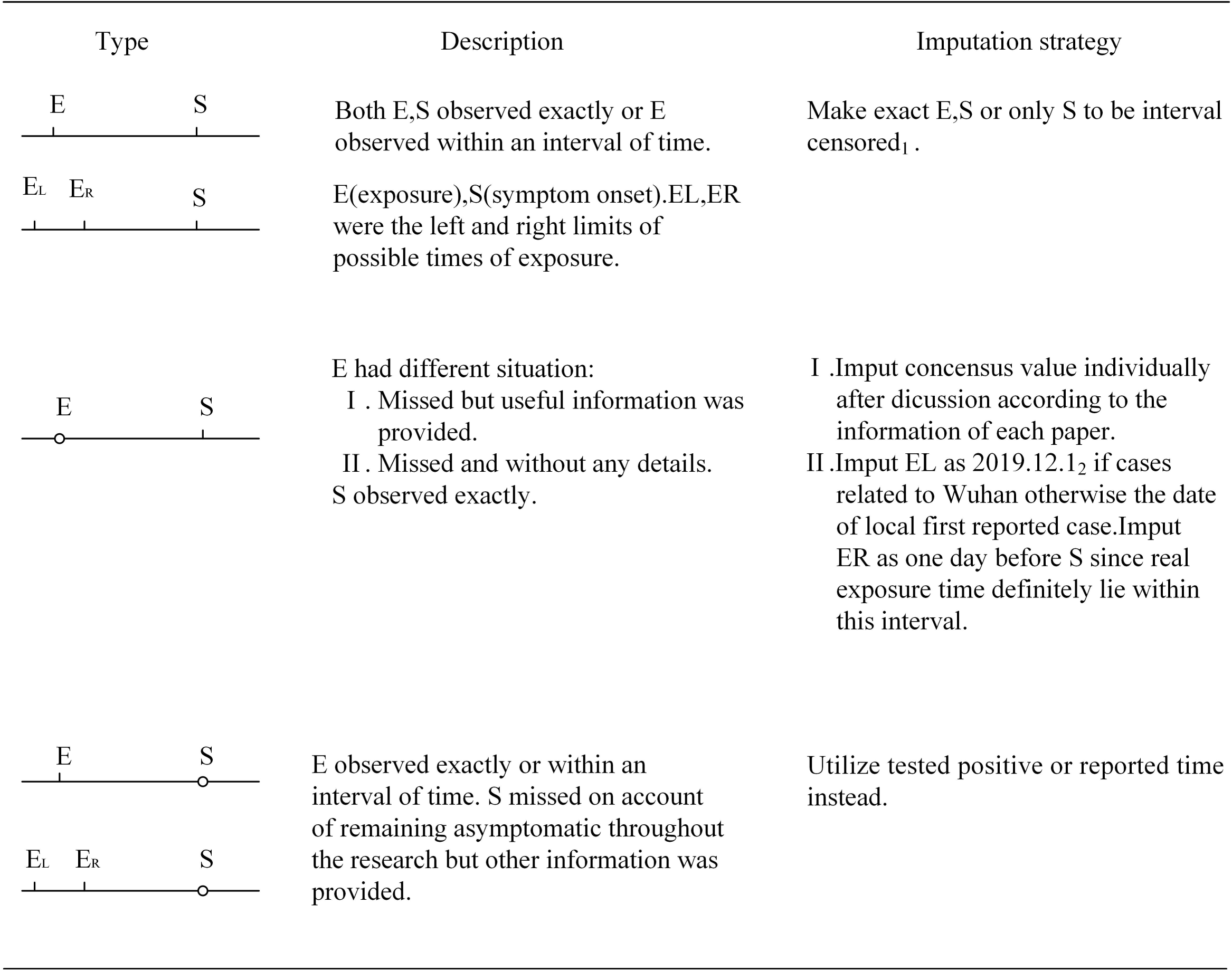
Data classification and imputing strategy.All types can be generally divided to three categories: non-missing data, exposure missed data and symptom onset missed data.

### Statistical analysis

Not all searched and collected documents had information on variation parameters, including standard deviations and variances (16). To ensure that all eligible studies were included and analyzed, we chose to intuitively show parameters, including ranges, confidence intervals (CIs), and central tendencies (medians or means) of studies in categories 1-3 rather than standard meta-analysis.

We performed pooled analysis with individual-level data extracted from studies in categories 3-4. We applied an accelerated failure time model to estimate the incubation period and its five important percentiles (2.5%, 25%, 50%, 75%, and 97.5%, respectively) with three distributions (log-normal, Weibull, and gamma) (17,18) and CIs by using bootstrapped and Markov chain Monte Carlo methods.

To examine the effect of our imputation strategy, we estimated the incubation period of merely non-missing data for sensitivity analysis. For the two types of asymptomatic patients, we considered all patients without symptoms in each study as truly asymptomatic and estimated the distribution of the time from exposure to the positive test. Between-study heterogeneity was also tested by using hierarchical models (16), which produced nearly identical results to the pooled analysis (not shown). All analyses were conducted using R software (v4.0.2, R Foundation; Vienna, Austria) with coarseDataTools, tidyverse, and lubridate packages.

## Results

We initially acquired 1071 studies from English and Chinese databases. After ruling out 190 duplicates, we screened titles and abstracts to exclude non-English, non-Chinese, and irrelevant studies. Finally, sixty-four articles from mainland China, Taiwan, and 8 other countries or regions were identified by 2 reviewers after reading full texts to determine whether they met the selection criteria (Figure 2). There were 30 studies with individual-level data from 945 cases, including 31 patients with incubation periods exceeding 14 days and 81 asymptomatic cases. Of these cases, 62% needed imputations (Table 1)and 854 cases were used for pooled analysis after excluding samples whose symptom onset preceded exposure (Table 2).

**Table 1:**
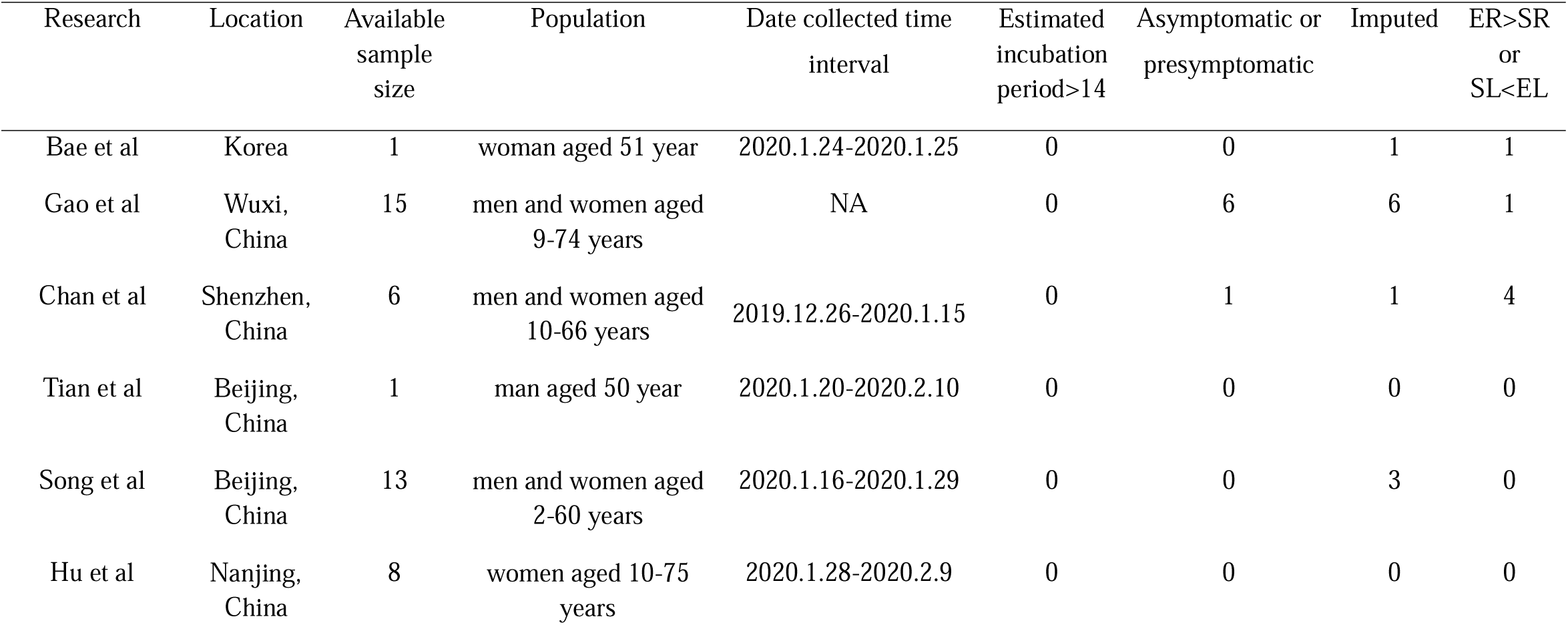
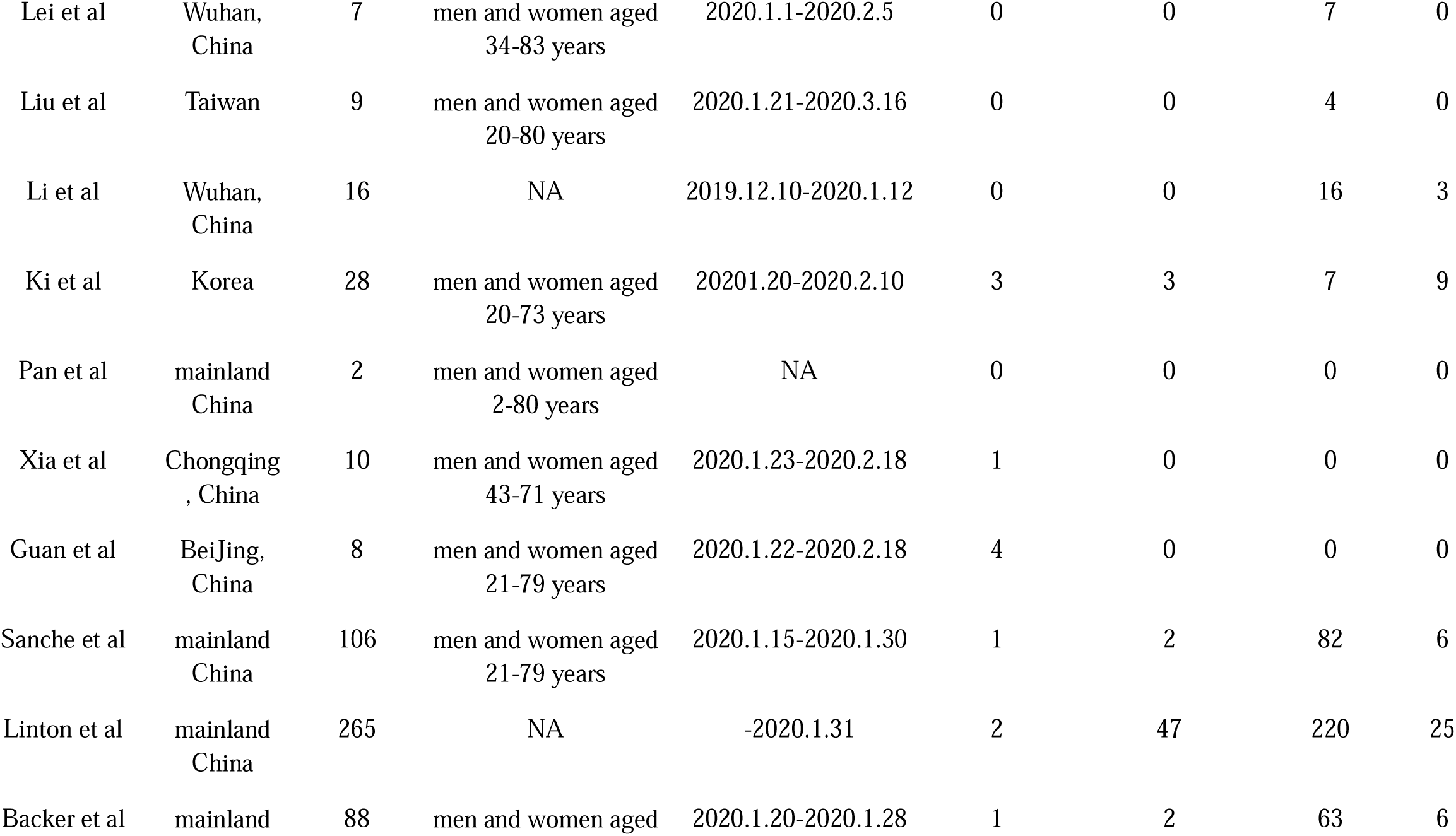
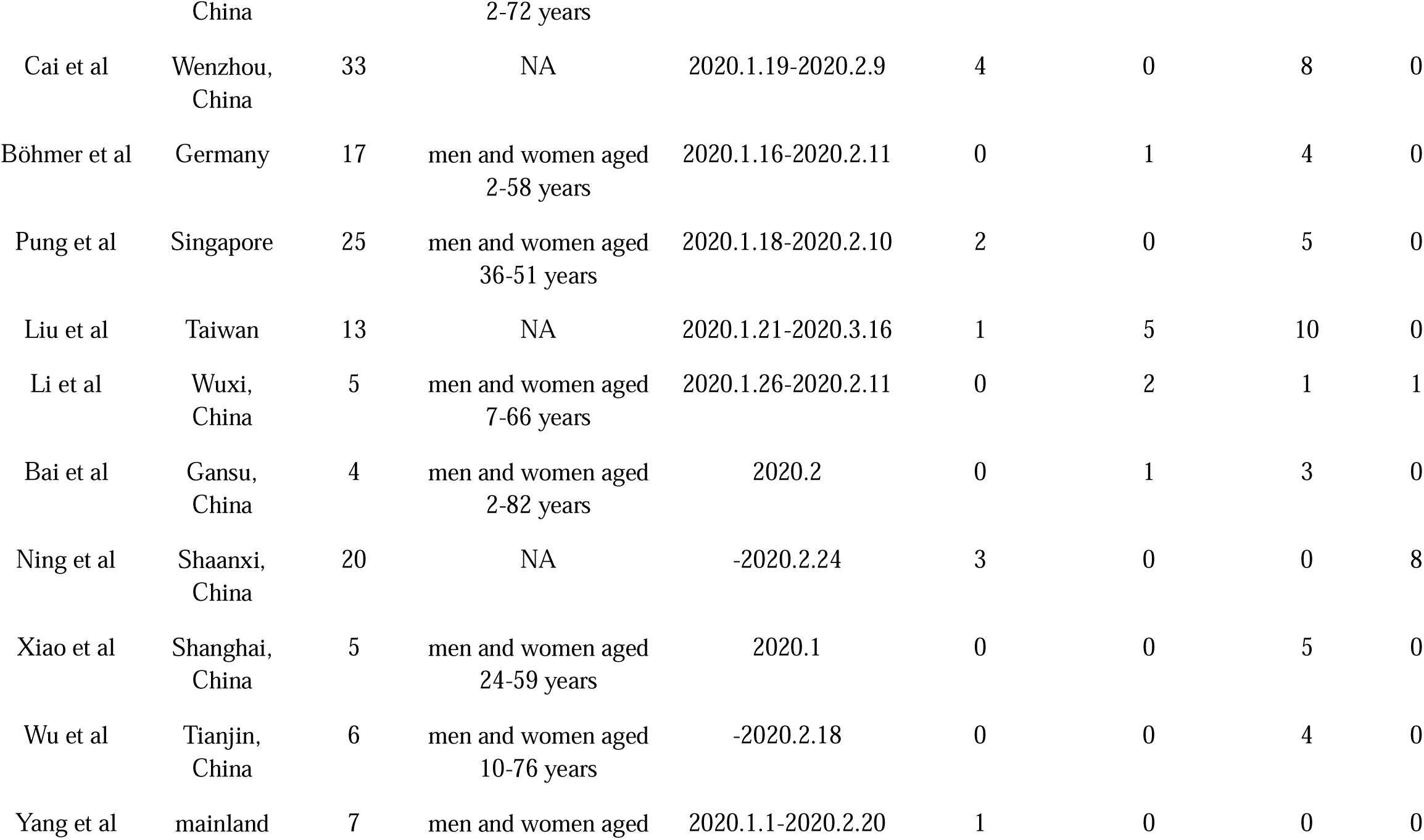
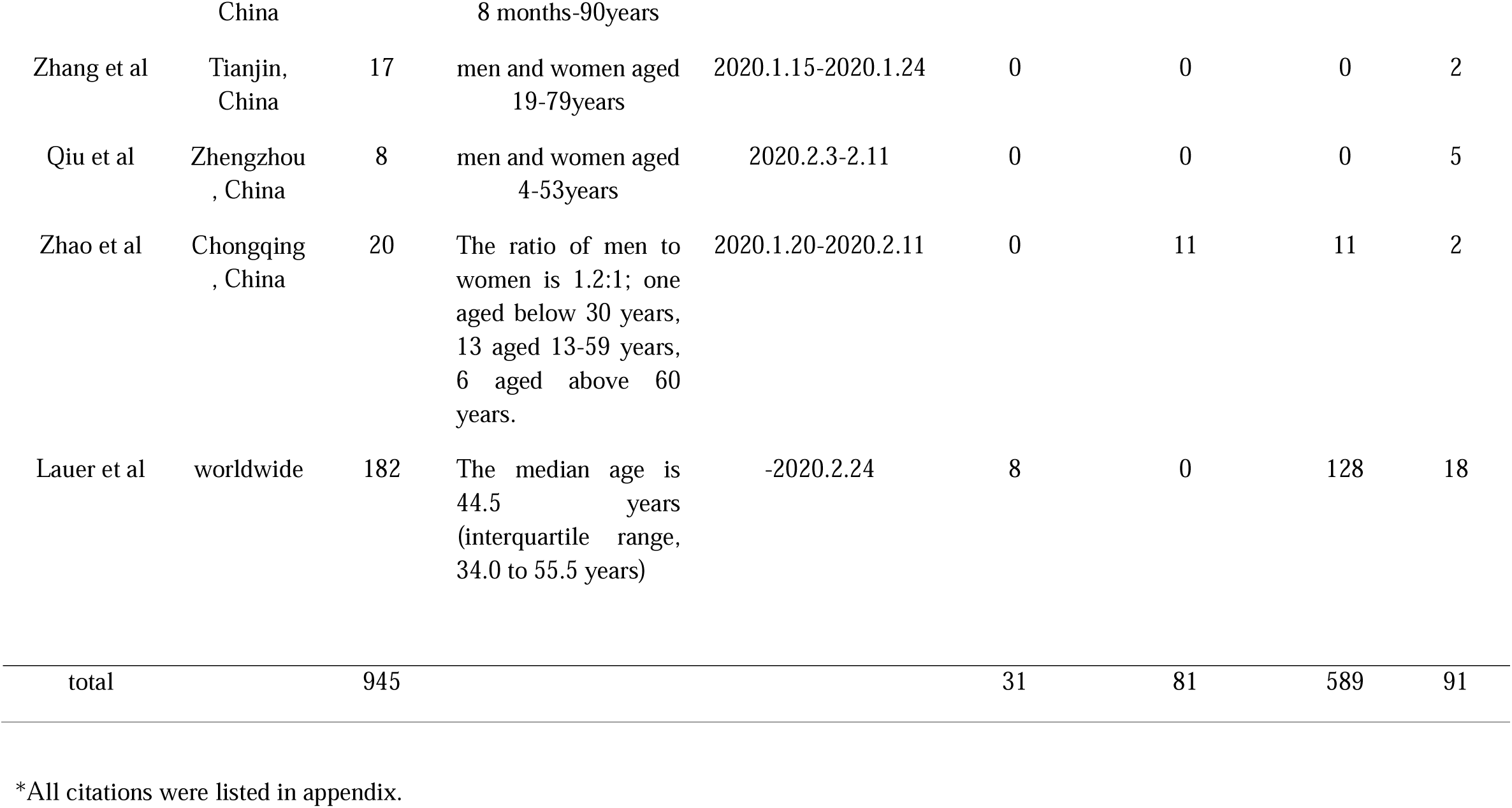
Studies included in pooled analysis*

**Table 2:**
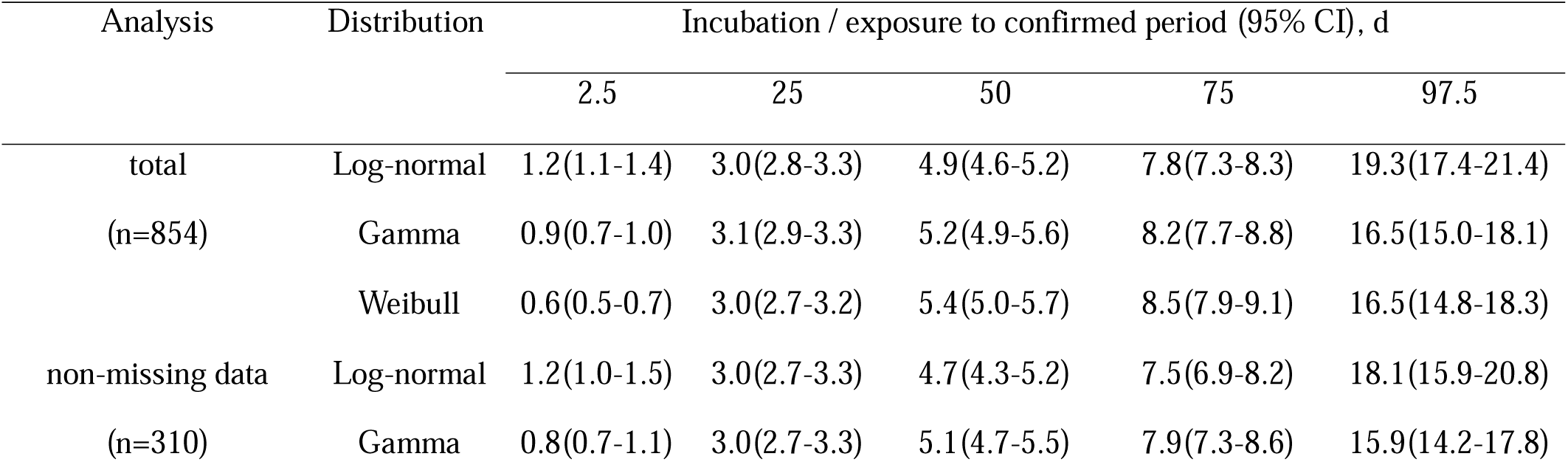
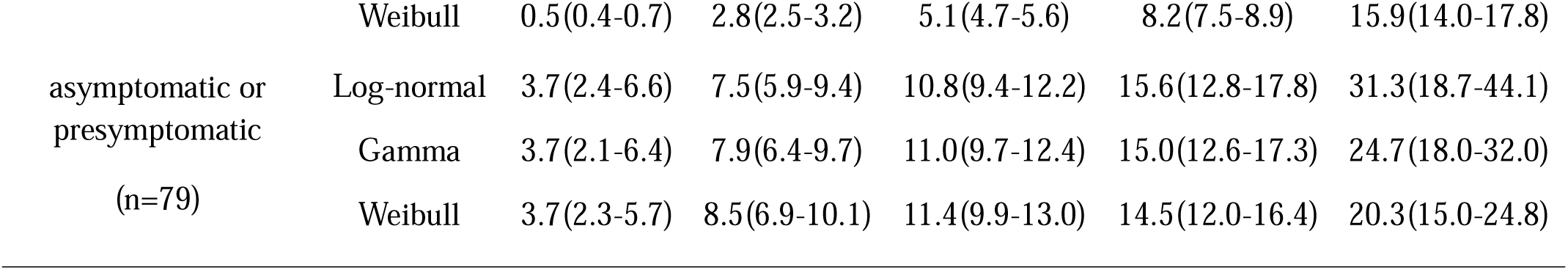
Five parameter estimates for three distributions based on total and part of 854 cases

**Figure 2:**
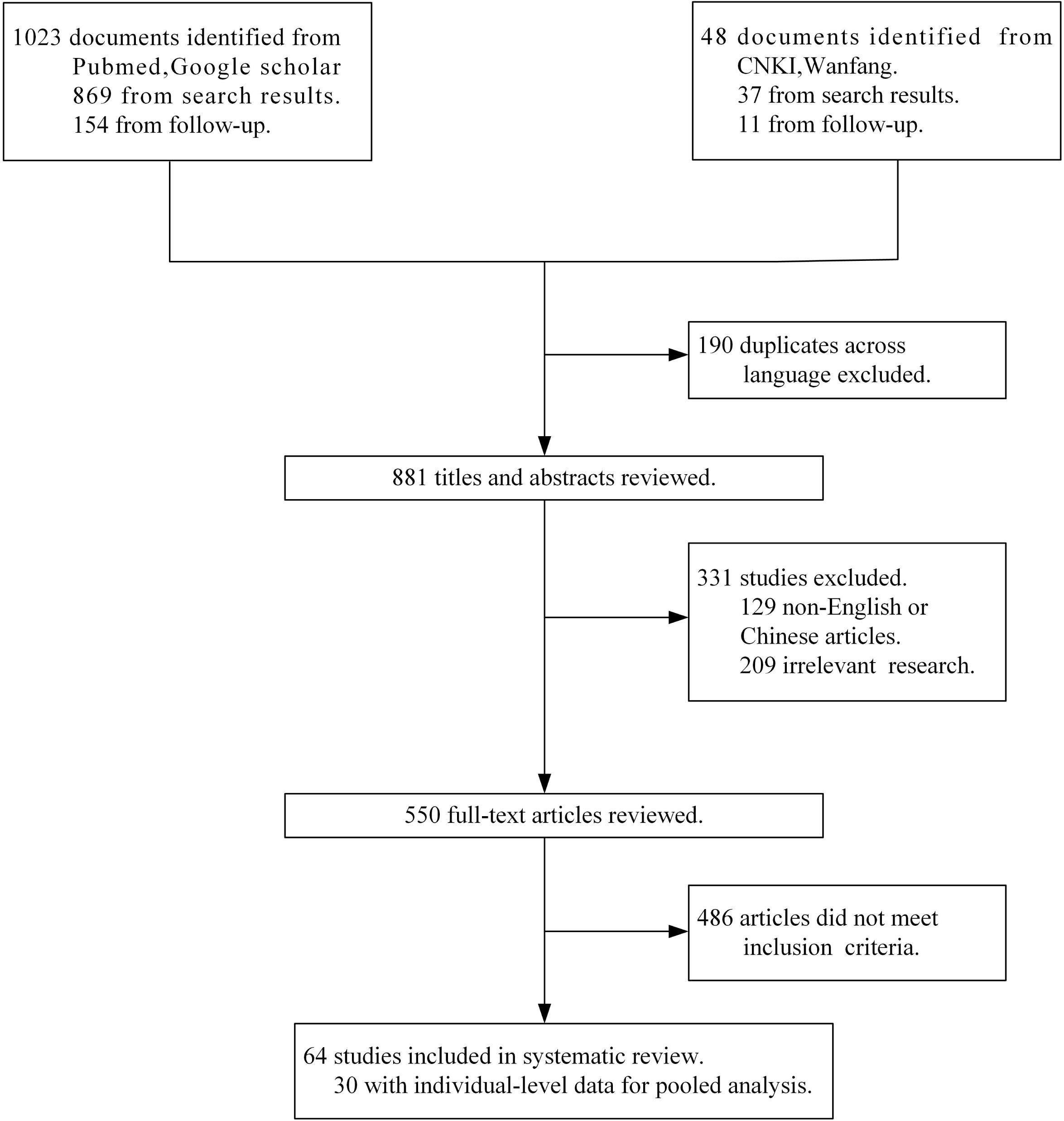
Literature search and selection

There were 58 central tendencies from 49 articles, including 6 articles with more than 1 central tendency. Of these 49 articles, 72% of their means and medians were distributed across 4-7 days and 26% of upper range limits and CIs exceeded 14 days (Figure 3). In the pooled analysis of 854 cases, we not only estimated the full distribution of incubation periods fitted in a log-normal distribution (Figure 4A), but also provided 5 important percentiles of all 3 distributions (Table 2). The estimated median incubation period from the log-normal distribution was 4.9 days (95% CI: 4.6-5.2 days) and the 97.5th percentile was 19.3 days (17.4-21.4 days). Approximately 6% of all cases had symptoms after 14 days.

**Figure 3:**
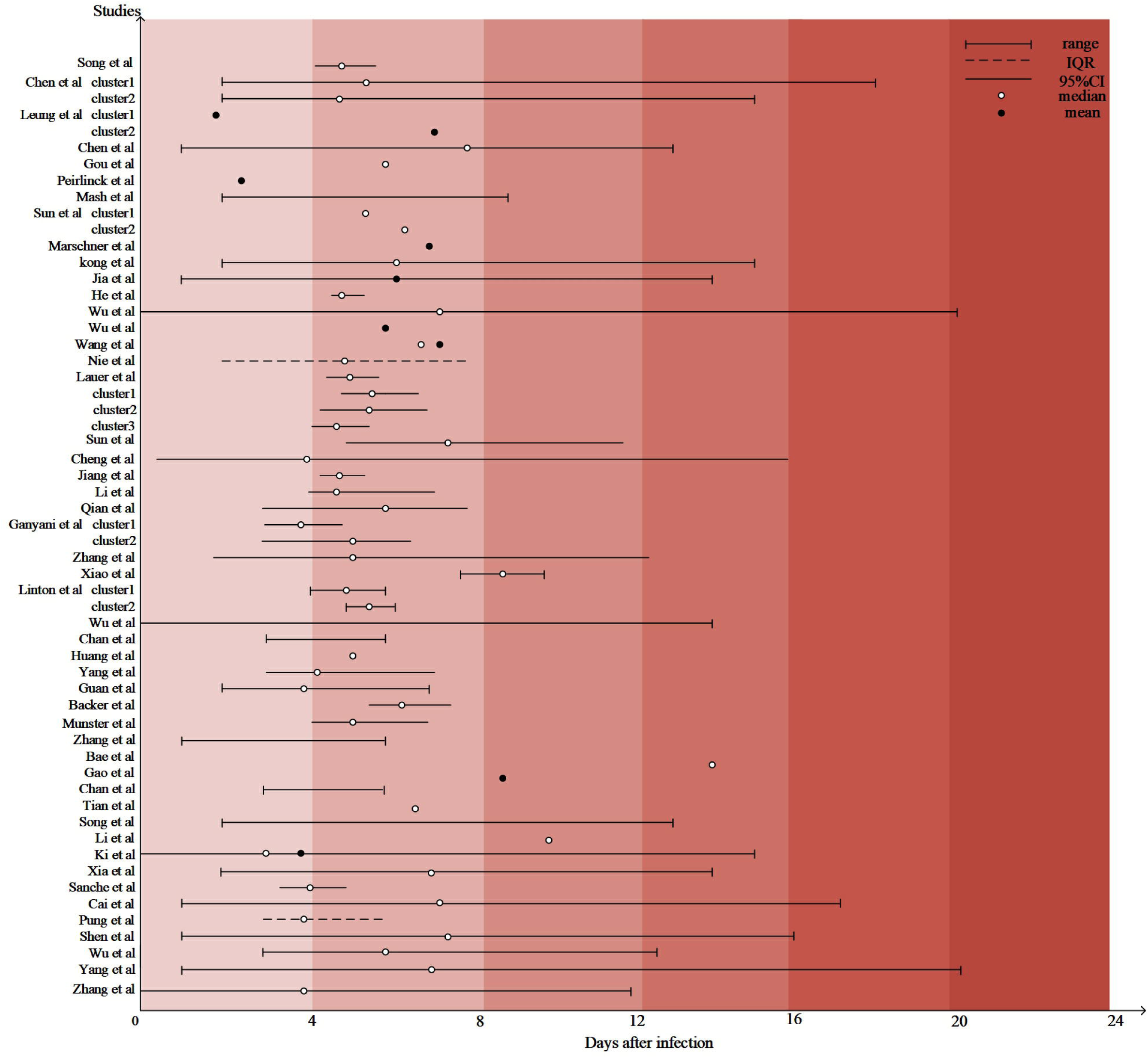
Visualization of 49 studies with central tendencies,ranges or confidential intervals.Studies that provided more than one estimates were performed with numbered clusters. All citations were listed in appendix.

**Figure 4:**
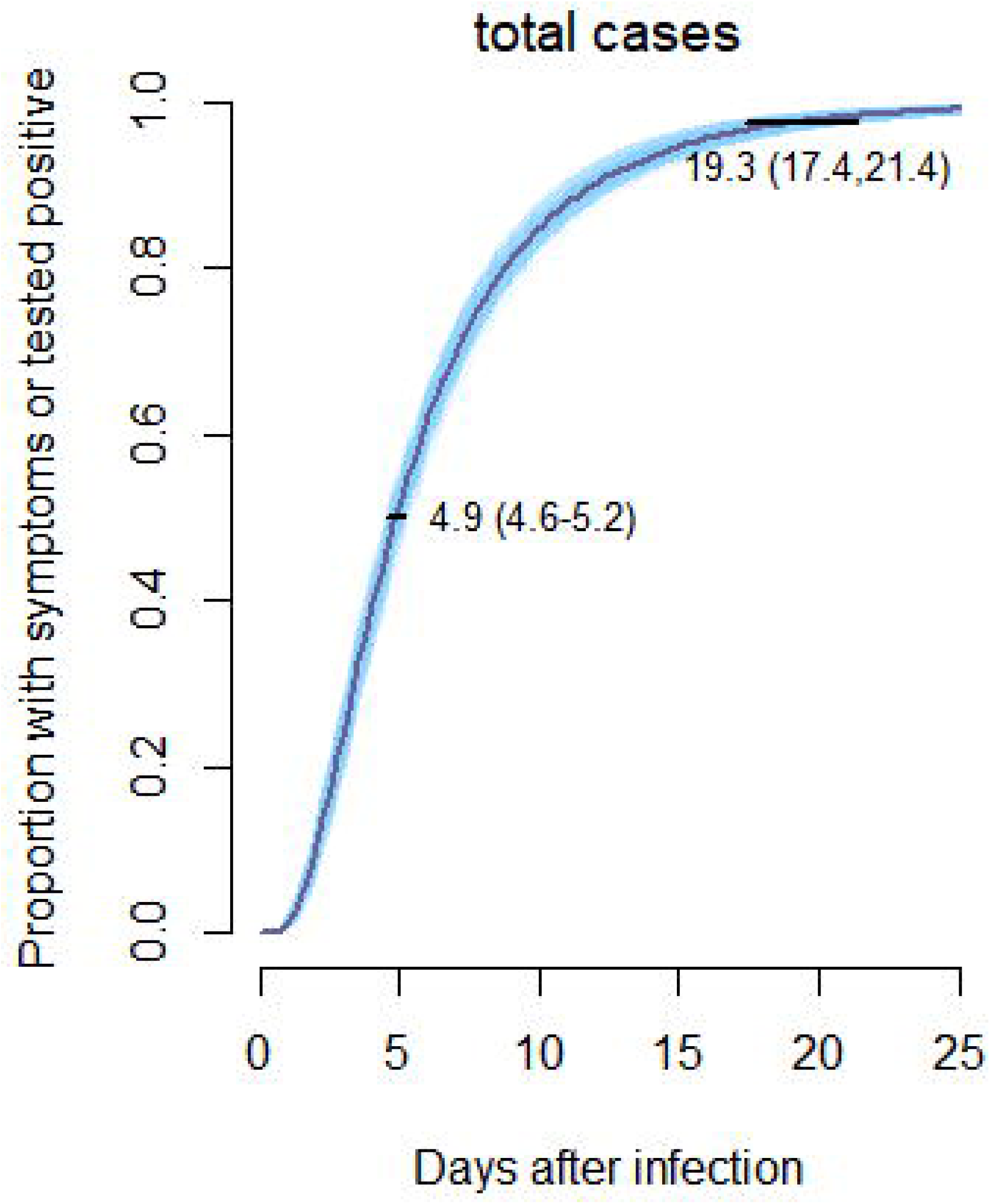
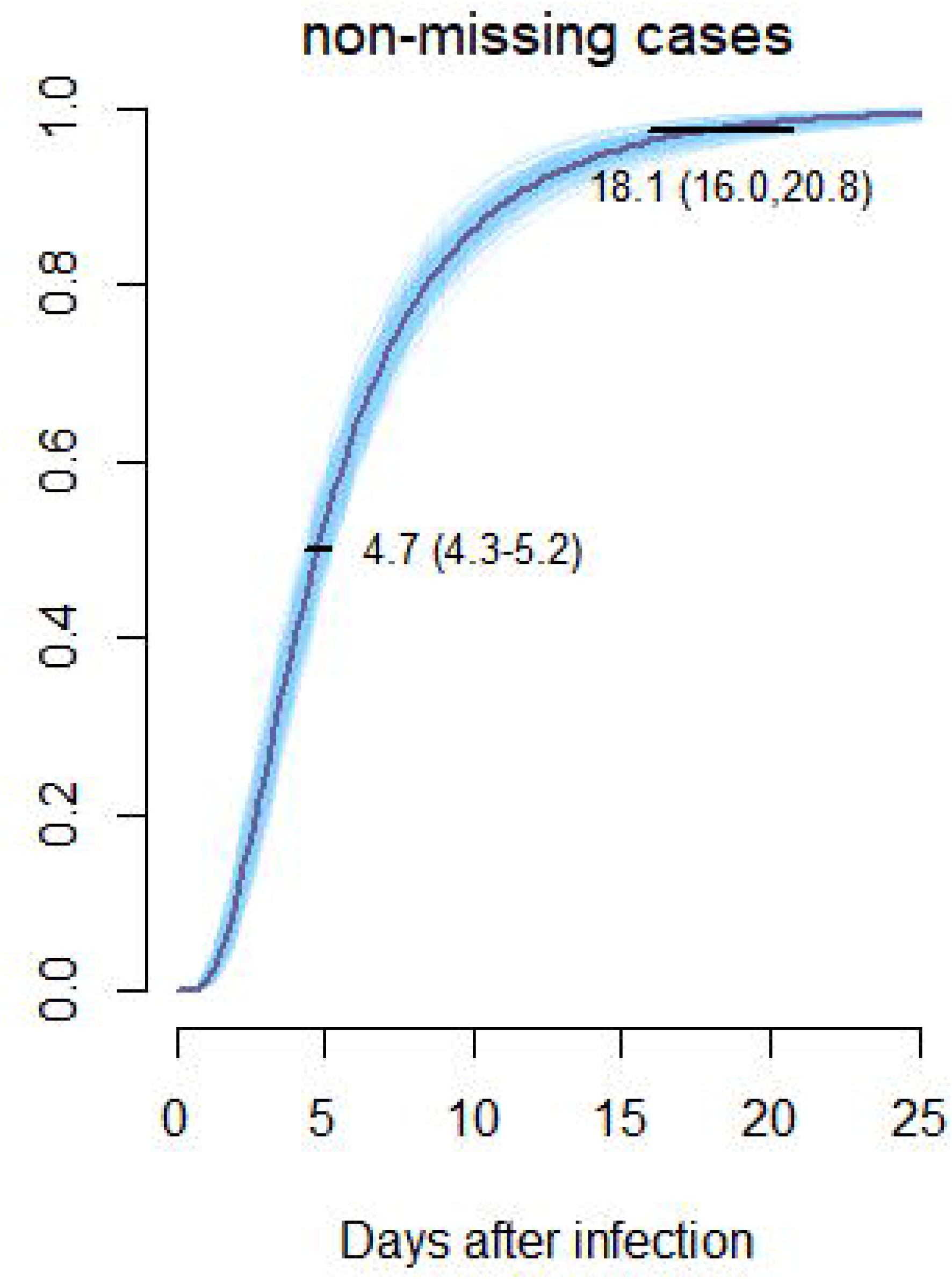
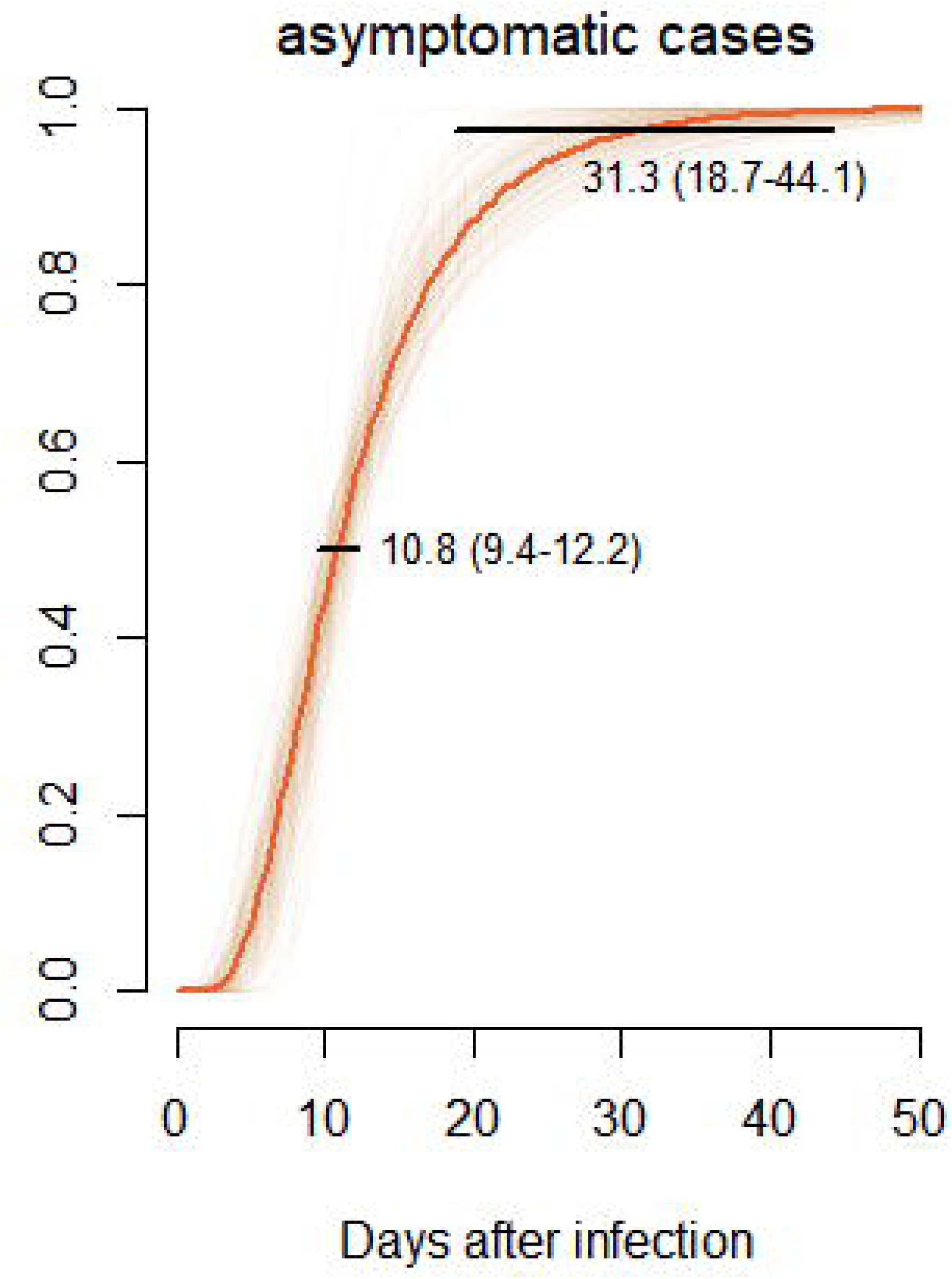
Estimating distributions of the incubation period and the time between exposure to tested positive.Using 854 cases (A),310cases(B) and 81cases(C) respectively.

To demonstrate the effectiveness of our imputation strategy, we estimated the distribution of incubation periods based on 310 cases with non-missing data in which the median was 4.7 days (95% CI: 4.3-5.2 days) and the 97.5th percentile was 18.1 days (16.0-20.8 days) (Figure 4B). Additionally, the distribution of time from exposure to a positive test was also provided, for which the median and 97.5th percentile were 10.8 days (95% CI: 9.4-12.2 days) and 31.3 days (18.7-44.1 days), respectively (Figure 4C).

## Discussion

In this study, we present both an intuitive visualization of complete incubation period-related studies and full distributions of time from exposure to symptom onset based on pooled analysis. Our estimated median incubation period was 4.9 days, which is generally consistent with the results of formal published studies (18), and close to the estimates for severe acute respiratory syndrome (SARS) and Middle East respiratory syndrome (MERS) (4.6 days and 5.2 days, respectively) (19). The 97.5th percentile was 19.3 days in our study, as we included more asymptomatic and presymptomatic patients extracted from studies without the tendency to bring in more patients with severe symptoms and a short incubation period.

When we imputed missing data cases, excluding Wuhan-related cases, we used the date of the first locally reported patient as EL, while the actual exposure might have been earlier than this. Thus, we imputed EL as December 1, 2019, the date of symptom onset of the first known case (1) for all exposure-missing cases, and there were no obvious differences compared to our original imputation method (4). A sensitivity analysis that estimated the incubation period based on mere non-missing data was performed to examine our entire imputation strategy, and the results were consistent with our estimates based on total data. Since the heterogeneity between studies can be misleadingly large when quantified by I^2^ of observational studies (20,21), we used hierarchical models instead which showed similar results to pooled analysis (not shown).

To ensure the comprehensiveness of our included studies, we did not use the standard meta-analysis method that acquires variation parameters; thus, we abandoned generating combined results from different studies. In addition, like those from Chinese researchers, there were a number of studies published only in languages other than English or Chinese, but these articles were not included because of the limits of languages. Moreover, compared to the incubation period estimates based on the data that imputed the positive test date as the date of symptom onset, we believe that the actual incubation period probably exceeds this because the presymptomatic patients included will eventually develop symptoms.

The current 14-day isolation period is mainly based on previous COVID-19 studies conducted in the early stage of the outbreak. Along with the global spread of the virus, cases of both types of asymptomatic COVID-19 cases gradually increased. On the contrary, we were not aware of any reviews or pooled analyses of incubation period estimates based on all kinds of infections. According to our research and that of Kimball et al. (7), Wu et al. (8), and Borras et al. (22), only 94% of cases will have symptoms during isolation if the current 14-day quarantine strategy is maintained, and >10% of total patients are asymptomatic, which can infect other healthy individuals if the rules of quarantine are such that cases are released from isolation based merely on whether they have symptoms.

In China, the government ruled that patients without symptoms can only be released after they have tested negative twice (23). Such policies require abundant resources for both medical and public health. In addition, trying to trace and identify asymptomatic individuals can be costly to countries still in an outbreak, especially in underdeveloped areas. Since testing each individual is still unachievable for now, in addition to imperfections of detection tools like false-negative results, properly prolonging the isolation or quarantine period would contribute to identifying patients with longer incubation periods and those without any symptoms. Thus, we can separate more susceptible populations from infected ones, which will not only help control the outbreak, but will also further reduce sporadic cases.

## Data Availability

All data are available from authors on request

## Acknowledgments

The authors would like to thank the Shanxi health commission for the grant of the special foundation of COVID-19(No.16). We also appreciate researchers who generously shared data in their studies.

## Author Bio

A postgraduate of Shanxi medical university, mainly focus on statistical theories and methods applied in the epidemiology of both infectious and chronic diseases.

## References

1. Li, Q., Guan, X., Wu, P., Wang, X., Zhou, L., Tong, Y., Ren, R., Leung, K. S. M., Lau, E. H. Y., Wong, J. Y., Xing, X., Xiang, N., Wu, Y., Li, C., Chen, Q., Li, D., Liu, T., Zhao, J., Liu, M., … Feng, Z. (2020). Early Transmission Dynamics in Wuhan, China, of Novel Coronavirus-Infected Pneumonia. The New England Journal of Medicine, 382(13), 1199–1207. https://doi.org/10.1056/NEJMoa2001316

2. World Health Organization. Coronavirus disease 2019 (COVID-19): Situation Report – 175. 13 July 2020. Accessed https://www.who.int/docs/default-source/coronaviruse/situation-reports/20200713-COVID-19-sitrep-175.pdf?sfvrsn=d6acef25_2

3. Nelson KE, Williams CM. Infectious disease epidemiology, theory and practice. Sudbury, MA: Jones & Bartlett Publishers, 2006.

4. Lauer SA, Grantz KH, Bi Q, et al. Estimating the incubation time of the novel coronavirus (COVID-19) based on publicly reported cases using coarse data tools. 2020. Accessed at https://github.com/HopkinsIDD/ncov_incubation on 3 March 2020

5. Bureau of Disease Control and Prevention, National Health Commission of the People’s Republic of China. Protocol on the prevention and control of Novel Coronavirus Pneumonia [Edition7]. http://www.nhc.gov.cn/yzygj/s7653p/202003/46c9294a7dfe4cef80dc7f5912eb1989.shtml

6. TheWhite House. Press Briefing by Members of the President’s Coronavirus Task Force. 31 January 2020. Accessed at www.whitehouse.gov/briefings-statements/press-briefing-members-presidents-coronavirus-task-force on 1 February 2020

7. Kimball A, Hatfield KM, Arons M, et al. Asymptomatic and Presymptomatic SARS-CoV-2 Infections in Residents of a Long-Term Care Skilled Nursing Facility - King County, Washington, March 2020. MMWR Morb Mortal Wkly Rep. 2020;69(13):377-381. Published 2020 Apr 3. doi:10.15585/mmwr.mm6913e1

8. Wu ZY. Contribution of asymptomatic and pre-symptomatic cases of COVID-19 in spreading virus and targeted control strategies. Zhonghua Liu Xing Bing Xue Za Zhi. 2020;41(6):801–805..doi:10.3760/cma.j.cn112338-20200406-00517

9. He X, Lau EHY, Wu P, et al. Temporal dynamics in viral shedding and transmissibility of COVID-19. Nat Med. 2020;26(5):672–675. doi:10.1038/s41591-020-0869-5

10. Guan WJ, Ni ZY, Hu Y, Liang WH, Ou CQ, He JX, et al. China Medical Treatment Expert Group for COVID-19. Clinical characteristics of coronavirus disease 2019 in China. N Engl J Med. 2020;NEJMoa2002032;

11. Ahrens W, Pigeot I, eds. Handbook of epidemiology. Berlin: Springer-Verlag, 2005.

12. Lauer SA, Grantz KH, Bi Q, et al. The Incubation Period of Coronavirus Disease 2019 (COVID-19) From Publicly Reported Confirmed Cases: Estimation and Application. Ann Intern Med. 2020;172(9):577–582. doi:10.7326/M20-0504

13. Linton, N. M., Kobayashi, T., Yang, Y., Hayashi, K., Akhmetzhanov, A. R., Jung, S.-M., Yuan, B., Kinoshita, R., & Nishiura, H. (2020). Incubation Period and Other Epidemiological Characteristics of 2019 Novel Coronavirus Infections with Right Truncation: A Statistical Analysis of Publicly Available Case Data. Journal of Clinical Medicine, 9(2). https://doi.org/10.3390/jcm9020538

14. Majumder MS, Mandl KD. Early in the epidemic: impact of preprints on global discourse about COVID-19 transmissibility. Lancet Glob Health. 2020;8(5):e627–e630. doi:10.1016/S2214-109X(20)30113-3

15. Reich NG, Lessler J, Cummings DA, Brookmeyer R. Estimating incubation period distributions with coarse data. Stat Med. 2009;28(22):2769–2784.

16. Lessler J, Reich NG, Brookmeyer R, Perl TM, Nelson KE, Cummings DA. Incubation periods of acute respiratory viral infections: a systematic review. Lancet Infect Dis. 2009;9(5):291–300. doi:10.1016/S1473-3099(09)70069-6

17. Sartwell PE. The distribution of incubation periods of infectious disease. Am J Hyg 1950; 51:310–8.

18. Cowling BJ, Muller MP, Wong IOL, Ho L, Louie M, McGeer A, Leung G. Alternative methods of estimating an incubation distribution: examples from severe acute respiratory syndrome. Epidemiology 2007; 18(2):253–9.

19. Zumla A, Hui DS, Perlman S. Middle East respiratory syndrome. Lancet. 2015;386(9997):995–1007. doi:10.1016/S0140-6736(15)60454-8

20. Guyatt GH, Oxman AD, Kunz R, et al. GRADE guidelines, 7: rating the quality of evidence—inconsistency. J Clin Epidemiol 2011; 64: 1294–302.

21. Iorio A, Spencer FA, Falavigna M, et al. Use of GRADE for assessment of evidence about prognosis: rating confidence in estimates of event rates in broad categories of patients. BMJ 2015; 350: h870

22. Borras-Bermejo B, Martínez-Gómez X, San Miguel MG, et al. Asymptomatic SARS-CoV-2 Infection in Nursing Homes, Barcelona, Spain, April 2020 [published online ahead of print, 2020 Jun 23]. Emerg Infect Dis.

23. Wu J, Lu AD, Zhang LP, Zuo YX, Jia YP. Zhonghua Xue Ye Xue Za Zhi. 2019;40(1):52–57. doi:10.3760/cma.j.issn.0253-2727.2019.01.010

